# Seroprevalence of Antibodies to SARS-CoV-2 in Six Sites in the United States, March 23-May 3, 2020

**DOI:** 10.1101/2020.06.25.20140384

**Authors:** Fiona P. Havers, Carrie Reed, Travis Lim, Joel M. Montgomery, John D. Klena, Aron J. Hall, Alicia M. Fry, Deborah L. Cannon, Cheng-Feng Chiang, Aridth Gibbons, Inna Krapiunaya, Maria Morales-Betoulle, Katherine Roguski, Mohammad Rasheed, Brandi Freeman, Sandra Lester, Lisa Mills, Darin S. Carroll, S. Michele Owen, Jeffrey A. Johnson, Vera Semenova, State Collaborator Group, Jarad Schiffer, Natalie Thornburg, Carina Blackmore, Debra Blog, Angela Dunn, Scott Lindquist, Scott Pritchard, Lynn Sosa, George Turabelidze, John Wiesman, Randall W. Williams

## Abstract

**Importance:** Reported cases of SARS-CoV-2 infection likely underestimate the prevalence of infection in affected communities. Large-scale seroprevalence studies provide better estimates of the proportion of the population previously infected.

**Objective:** To estimate prevalence of SARS-CoV-2 antibodies in convenience samples from several geographic sites in the United States.

**Design:** Serologic testing of convenience samples using residual sera obtained for routine clinical testing by two commercial laboratory companies.

**Setting:** Connecticut (CT), south Florida (FL), Missouri (MO), New York City metro region (NYC), Utah (UT), and Washington State’s (WA) Puget Sound region.

**Participants:** Persons of all ages with serum collected during intervals from March 23 through May 3, 2020.

**Exposure:** SARS-CoV-2 virus infection.

**Main outcomes and measures:** We estimated the presence of antibodies to SARS-CoV-2 spike protein using an ELISA assay. We standardized estimates to the site populations by age and sex. Estimates were adjusted for test performance characteristics (96.0% sensitivity and 99.3% specificity). We estimated the number of infections in each site by extrapolating seroprevalence to site populations. We compared estimated infections to number of reported COVID-19 cases as of last specimen collection date.

**Results:** We tested sera from 11,933 persons. Adjusted estimates of the proportion of persons seroreactive to the SARS-CoV-2 spike protein ranged from 1.13% (95% confidence interval [CI] 0.70-1.94) in WA to 6.93% (95% CI 5.02-8.92) in NYC (collected March 23-April 1). For sites with later collection dates, estimates ranged from 1.85% (95% CI 1.00-3.23, collected April 6-10) for FL to 4.94% (95% CI 3.61-6.52) for CT (April 26-May 3). The estimated number of infections ranged from 6 to 24 times the number of reported cases in each site.

**Conclusions and relevance:** Our seroprevalence estimates suggest that for five of six U.S. sites, from late March to early May 2020, >10 times more SARS-CoV-2 infections occurred than the number of reported cases. Seroprevalence and under-ascertainment varied by site and specimen collection period. Most specimens from each site had no evidence of antibody to SARS-CoV-2. Tracking population seroprevalence serially, in a variety of specific geographic sites, will inform models of transmission dynamics and guide future community-wide public health measures.

**Key findings:** *Question:* What proportion of persons in six U.S. sites had detectable antibodies to SARS-CoV-2, March 23-May 3, 2020?

*Findings:* We tested 11,933 residual clinical specimens. We estimate that from 1.1% of persons in the Puget Sound to 6.9% in New York City (collected March 23-April 1) had detectable antibodies. Estimates ranged from 1.9% in south Florida to 4.9% in Connecticut with specimens collected during intervals from April 6-May 3. Six to 24 times more infections were estimated per site with seroprevalence than with case report data.

*Meaning:* For most sites, evidence suggests >10 times more SARS-CoV-2 infections occurred than reported cases. Most persons in each site likely had no detectable SARS-CoV-2 antibodies.

## Introduction

The first case of SARS-CoV-2 infection in the United States was reported in Washington State on January 20, 2020. The first U.S. case linked to community transmission was reported in California on February 26, 2020, followed by subsequent cases resulting from community transmission reported in Washington on February 28 and New York on March 3.^1-5^ Since January 2020, states have been recommended to report all laboratory-confirmed cases to CDC.^6^ However, reported cases likely represent only a fraction of all SARS-CoV-2 infections, as an unknown proportion of cases are mild or asymptomatic, or otherwise not diagnosed or ascertained through passive public health reporting.^7-9^ Furthermore, viral testing has been limited in many sites, was often reserved for severely ill patients early in the U.S. outbreak, and testing availability has changed rapidly. Each of these issues could have confounded estimates of incident cases and epidemic dynamics that use only case-based reporting data.

Detection of antibodies to SARS-CoV-2 in a person’s blood likely indicates they were infected at some point since the start of the pandemic. Thus, serologic assays can be used to determine population-based estimates of infection, including those who had mild or asymptomatic infection or who were never tested despite symptoms.

We used convenience samples of available residual clinical specimens obtained from two commercial diagnostic laboratories to conduct a serologic survey. Our goal was to estimate the seroprevalence in the population, or proportion of the population with evidence of previous infection with SARS-CoV-2, by age group, in six geographically diverse U.S. sites with known community transmission.

## Methods

We obtained convenience samples of residual patient sera collected for routine screening (e.g., cholesterol screening) or clinical management by two commercial clinical laboratories (Lab A and Lab B) from six sites collected during discrete periods from March 23 through May 3 (**Table 1, Figure 1)**. Sites included the Puget Sound region of Washington State (WA) (defined broadly); the New York City metro region (NY) (defined broadly); south Florida (FL) (restricted to Miami-Dade, Broward, Palm Beach and Martin counties); and all of Missouri (MO), Utah (UT), and Connecticut (CT) **(eFigure 1 a-f)**. Age or age group, patient sex, and collection date were available for all specimens; we aimed to have at least 300 specimens per age group. Lab A specimens from NY, WA, and FL and Lab B specimens from MO, UT, and CT came from unique individuals; it was unknown whether multiple specimens came from the same individual for Lab B specimens from WA and NY. Zip code of patient residence was known for Lab A specimens and all Lab B specimens except those from NY and WA. Based on information from Lab B, which indicated that the majority of specimens from its facilities in WA and NY were drawn from the areas of greatest population density in Puget Sound and NYC metropolitan regions, respectively, we assumed that Lab B specimens from WA and NY were from a similar geographic distribution to those received from Lab A for those sites geographic distribution. No information on the reason for specimen collection was available. This protocol underwent review by CDC human subjects research officials, who determined that the testing represented non-research activity in the setting of a public health response to the COVID-19 pandemic.

**Table 1.** Number of residual clinical specimens from commercial laboratories tested for antibodies to SARS-CoV-2 in six geographic sites, by sex, age group, and location, with dates of specimen collection for each site.

|  | **Washington**<br>**(Puget Sound**<br>**Region)** | **New York City**<br>**Metro Region** | **South Florida** | **Missouri** | **Utah** | **Connecticut** |
| --- | --- | --- | --- | --- | --- | --- |
| **Date of**<br>**specimen**<br>**collection** | 3/23/20 – 4/1/20 | 3/23/20 – 4/1/20 | 4/6/20 – 4/10/20 | 4/20/20 –<br>4/26/20 | 4/20/20 – 5/3/20 | 4/26/20 – 5/3/20 |
| **Sex** |  |  |  |  |  |  |
| **Female** | 1930 (59.1%) | 1333 (53.71%) | 964 (55.3%) | 1018 (54.09%) | 673 (59.1%) | 729 (50.94%) |
| **Male** | 1334 (40.9%) | 1149 (46.29%) | 778 (44.7%) | 864 (45.91%) | 465 (40.9%) | 702 (49.06%) |
| **Age group** |  |  |  |  |  |  |
| **0-18** | 219 (6.7%) | 311 (12.5%) | 69 (4.0%) | 158 (8.4%) | 25 (2.2%)<sup>a</sup> | 219 (15.3%) |
| **19-49** | 1213 (37.2%) | 909 (36.6%) | 491 (28.2%) | 394 (20.9%) | 470 (41.3%) | 297 (20.8%) |
| **50-64** | 782 (24.0%) | 455 (18.3%) | 326 (18.7%) | 405 (21.5%) | 328 (28.8%) | 300 (21.0%) |
| **65+** | 1050 (32.2%) | 807 (32.5%) | 856 (49.1%) | 925 (49.1%) | 315 (27.7%) | 615 (43.0%) |
| **TOTAL** | 3264 | 2482 | 1742 | 1882 | 1132 | 1431 |

<sup>a</sup> Excluded from analysis due to small sample size.

**Figure 1.**
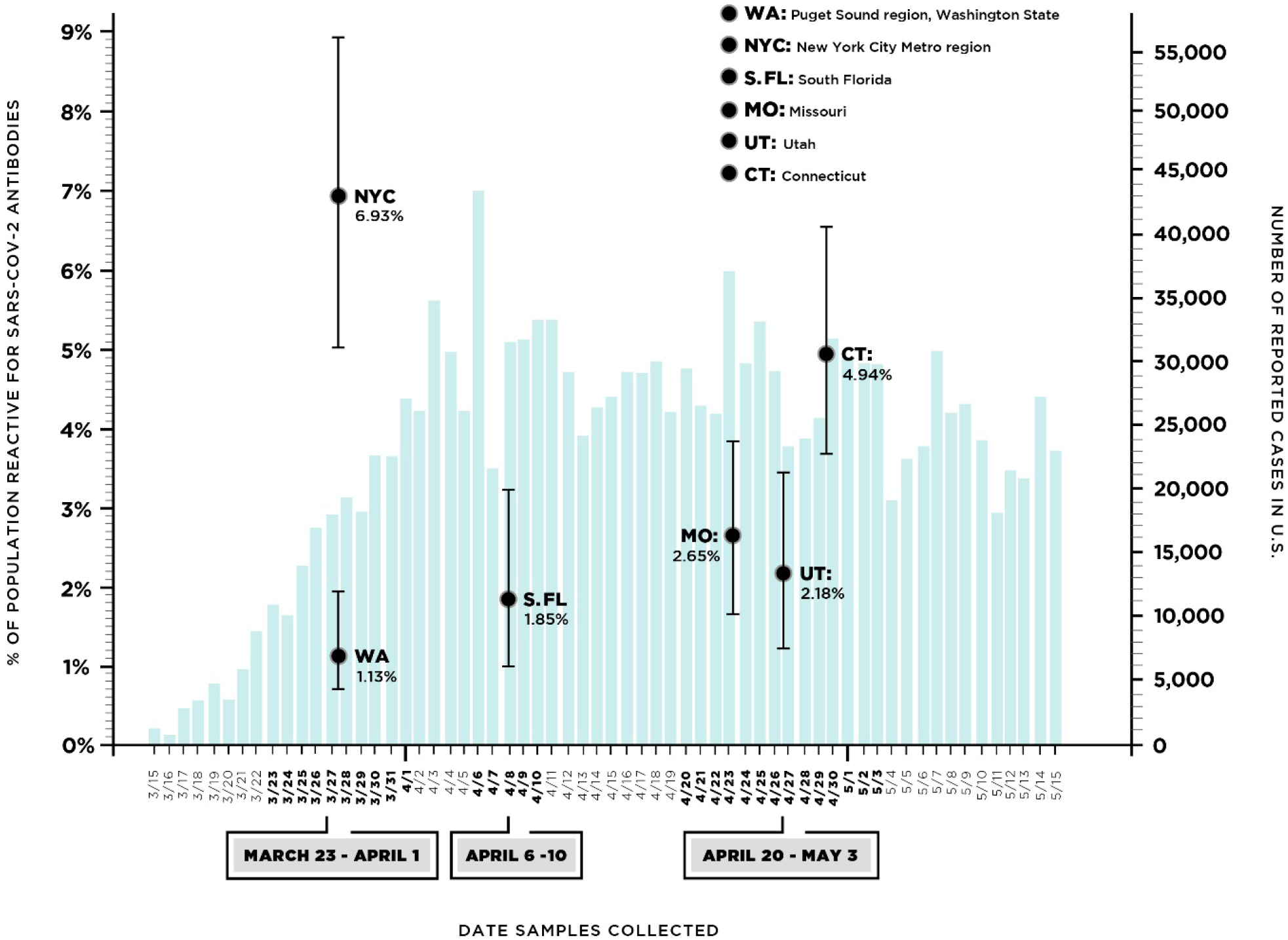
Timeline of reported cases in US, with specimen collection dates and estimates of seroprevalence to SARS-CoV-2 antibodies with 95% confidence intervals for six geographic sites from which residual clinical specimens were collected. Seroprevalence estimate is shown at the midpoint of the specimen collection date range.

### Laboratory methods

Sera were tested at CDC in a two-step process, a screening assay followed by a confirmatory assay for presumptive reactive specimens identified through screening. The CDC developed and validated enzyme-linked immunosorbent assay (ELISA) that was used as the confirmatory assay has been previously described.^10^ A specimen was considered reactive if, on confirmatory testing, at a background corrected optical density (OD) of 0.4 and at a serum dilution of 1:100, it had a signal to threshold ratio of >1. The screening assay was similar to the confirmatory assay. Sera were screened at a 1:100 dilution using a qualitative pan immunoglobulin (Ig) ELISA against the pre-fusion stabilized ectodomain of the SARS-CoV-2 spike protein (S).^11^ However, a greater coating concentration of spike protein was used, only 1 dilution was tested for each serum sample (1:100), and different OD cutoffs were first used to identify presumptive reactive specimens, which were then referred for confirmatory testing.^10^ Using the above definition of reactivity, specificity was 99.3% (confidence interval [CI] 98.32 – 99.88%) and sensitivity was 96.0% (CI 89.98 – 98.89%).^10^ Results of testing against sera from polymerase chain reaction– confirmed infections with other coronaviruses indicate that antibodies to commonly circulating human coronaviruses did exhibit some cross-reactivity, but this was below the limits of detection for this assay.^10^

### Analysis

We calculated seroprevalence as the proportion of specimens that were confirmed reactive, stratified by sex and age group (0 to ≤18 years, 19 to ≤49 years, 50 to ≤64 years, and 65+ years). We calculated age-and-sex-standardized seroprevalence estimates using weights derived from U.S. census county-level population projections for the most sampled counties for WA, NY, and FL, and from U.S. census state-level data for CT, MO, and UT. We estimated 95% CIs by generating 10,000 bootstrapped samples with replacement (**Supplement: Statistical Methods**). We then conducted additional analyses with bootstrapping to account for assay test performance, using the sensitivity and specificity parameters described above. We defined estimates that were age- and sex-standardized and adjusted for test characteristics as fully adjusted estimates. To assess for potential differences in populations using different laboratories, we compared seroprevalence in specimens from Lab A to those from Lab B for NY and WA, the two sites where both laboratories collected specimens.

To estimate the degree of under-ascertainment of reported cases for all sites, we assumed that the presence of SARS-CoV-2 antibodies represented infections that occurred prior to the last date of specimen collection. We applied the estimated age- and sex-adjusted seroprevalence estimates to the respective populations for each site to estimate total infections by that time point. We then divided these numbers by the cumulative number of official reported case counts reported to health departments^12^ as of the last date of specimen collection for each site. As antibodies may take an average of 10 to14 days to be detectable after infection,^13-15^ and collection periods were 6 to 14 days in length, we accounted for a lag in the development of antibodies in a scenario analysis using the cumulative number of reported cases as of 7 days prior to the start of specimen collection. The specimen collection period in relationship to reported cases by date is shown for each site in eFigure 2 (a-f).

R (Version 3.6.1) and Rstudio (version 1.2.1335) were used to perform statistical analysis.

## Results

We tested 11,933 residual sera specimens from six sites collected from March 23 through May 3, with discrete collection periods for each site within that timeframe **(Table 1)**. Among the 45.6% (1,488/3,264) Lab A specimens from WA for which zip-code data were available, 87.3% (1,299/1,488) came from King (n=786), Pierce (n=192), Snohomish (n=197), Kitsap (n=74) or Grays Harbor (n=50) counties; among the 27.4% (682/2,482) NYC specimens from Lab A with zip code data, 92.8% (633/682) came from Manhattan (n=243), Bronx (n=215), Brooklyn (n=104), Queens (n=56), or Nassau (n=15) counties. FL specimens were restricted to Miami-Dade (n=701), Broward (n=632), Palm Beach (n=376), and Martin (n=33) counties. Specimens were collected statewide from CT, MO, and UT, with most specimens collected from urban areas; location of specimen origin by zip code is shown in **eFigure 1 a-f**.

Seroprevalence estimates by sex and age, as well as fully adjusted estimates, are shown in **Table 2**. Seroprevalence ranged from 1.13% (95% CI 0.70 – 1.94) in WA to 6.93% (95% CI 5.02 – 8.92) in NYC for specimens collected March 23 – April 1. Seroprevalence estimates were in this range for the remaining four sites, which had later specimen collection dates. There was no clear trend by age and sex in these sites **(Figure 2)**. In NYC, there was a significant difference in fully adjusted seroprevalence between specimens obtained from Lab A (11.5%) and Lab B (5.70%; p <0.01). In WA, there was no difference in fully adjusted seroprevalence between specimens obtained from Lab A or B (1.86% v. 1.53%, p=0.47) (**eTable 1**).

**Table 2.** Percentage of reactive^a^ specimens, by sex and age, adjusted for assay performance characteristics^b^ for specimens collected in six geographic sites.

|  | % Reactive (95% confidence interval)<sup>a</sup> |  |  |  |  |  |
| --- | --- | --- | --- | --- | --- | --- |
|  | Washington<br>(Puget Sound<br>Region) | New York City<br>Metro Region | South Florida | Missouri | Utah | Connecticut |
| **Sex** |  |  |  |  |  |  |
| Male | 1.41 (0.76, 2.36) | 5.85 (4.46, 7.55) | 2.20 (1.09, 3.55) | 3.14 (1.83, 4.56) | 2.17 (0.85,3.94) | 5.65 (3.80, 7.60) |
| Female | 1.16 (0.67, 1.91) | 5.67 (4.20, 7.03) | 2.20 (1.23, 3.42) | 2.57 (1.48, 3.74) | 2.54 (1.21, 4.10) | 4.12 (2.55, 5.91) |
| **Age group, y** |  |  |  |  |  |  |
| 0-18 | 0.66 (0.0, 2.52)<sup>c</sup> | 2.74 (0.9, 5.03)<sup>c</sup> | 2.41 (0.00, 7.79) | 1.36 (0.00, 4.14) | --<sup>d</sup> | 0.81 (0.00, 2.89) |
| 19-49 | 1.32 (0.65, 2.27)<sup>c</sup> | 8.26 (6.18,<br>10.17)<sup>c</sup> | 0.87 (0.19, 2.22) | 3.36 (1.42, 5.53) | 1.82 (0.61, 3.52) | 6.08 (3.14, 9.29) |
| 50-64 | 0.85 (0.26, 1.94) | 6.52 (4.34, 9.61) | 1.95 (0.32, 4.00) | 1.96 (0.51, 3.76) | 2.89 (0.93, 5.21) | 8.11 (4.79, 11.64) |
| 65+ | 1.66 (0.85,2.74) | 3.66 (2.19, 5.19) | 3.04 (1.74, 4.45) | 3.23 (1.92, 4.58) | 2.70 (0.89, 5.04) | 4.15 (2.31, 6.04) |
| All ages, age-<br>standardized<sup>b</sup> | 1.13 (0.70, 1.94)<sup>e</sup> | 6.93 (5.02, 8.92)<sup>e</sup> | 1.85 (1.00, 3.23)<sup>e</sup> | 2.65 (1.65, 3.86)<sup>f</sup> | 2.18 (1.21, 3.43)<sup>f</sup> | 4.94 (3.61, 6.52)<sup>f</sup> |

<sup>a</sup> A specimen was considered reactive if, on confirmatory testing with the assay described in Freeman, et al.<sup>9</sup>, at a background corrected OD of 0.4, at a serum dilution of 1:100, it had a signal to threshold ratio of >1. <sup>b</sup> All estimates are adjusted for test performance characteristics (specificity 99.3% C) 98.32 – 99.88%; sensitivity 96.0% (CI 89.98 – 98.89%). <sup>c</sup> A subset of samples did not have age by year available and were instead classified as age 5-17, 18-49, 50-64 and 65+ years. The 5-17 and 19-49 age groups were combined with those 5-18 and 19-49 years, respectively. <sup>d</sup> The number of specimens for persons aged 0-18 years was inadequate, and those <18 years were excluded from the analysis. <sup>e</sup> Standardized to the age and sex distribution of the counties in each region from which most specimens originated, and adjusted for test performance characteristics as described above. <sup>f</sup> Standardized to the age and sex distribution of the state, and adjusted for test performance characteristics as described above.

**Figure 2.**
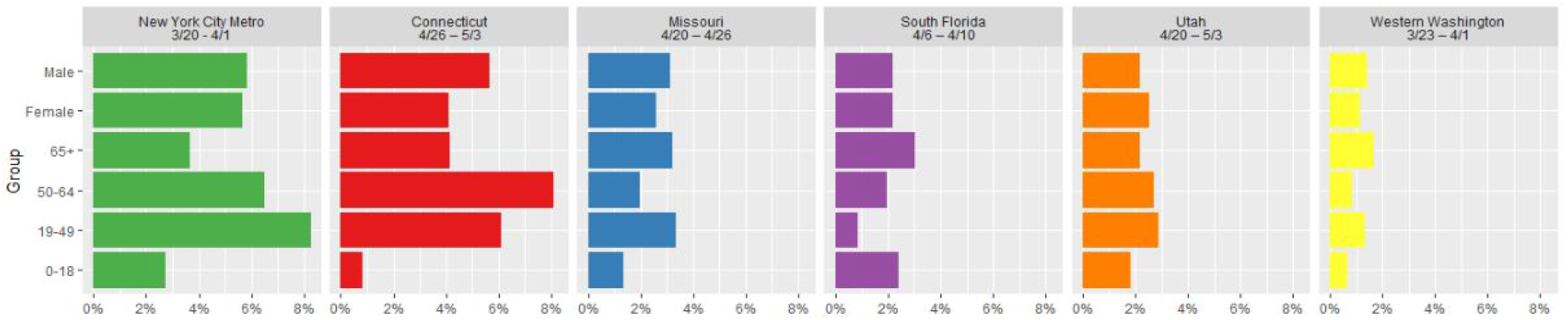
Estimates of seroprevalence to SARS-CoV-2 antibodies by age and sex in six geographic sites.

**Table 3** shows estimates of the number of SARS-CoV-2 infections suggested by seroprevalence estimates in each site and compares these to the number of reported cases as of the last date of specimen collection **(eFIgure 2 a-f)**. Our estimate for under-ascertainment was lowest in CT, where the estimated 176,012 infections were 6.0 (range 4.3 – 7.8) times greater than the 29,287 reported cases as of May 3, 2020, and highest for MO, where the 161,936 estimated infections were 23.8 (range 14.8 – 34.7) times greater than the 6,794 reported cases as of April 25, 2020. Estimated infections for all sites except CT were at least 10 times greater than the number of reported cases.

**Table 3.** Estimated number of infections based on seroprevalence estimates and comparison with the number of reported cases as of the last date of specimen collection for six sites.

|  | **Washington**<br>**(Puget Sound**<br>**Region)** | **New York City**<br>**Metro Region** | **South Florida** | **Missouri** | **Utah** | **Connecticut** |
| --- | --- | --- | --- | --- | --- | --- |
| Catchment description | King, Snohomish, Pierce, Kitsap and Grays Harbor | Manhattan, Bronx, Queens, Kings, Nassau | Miami-Dade, Broward, Palm Beach, Martin | State-wide | Adults 19+ (State-wide) | State-wide |
| Catchment population | 4,273,548 | 9,260,870 | 6,345,345 | 6,110,800 | 2,173,082 | 3,562,989 |
| Estimated seroprevalence<sup>a</sup> | 1.13% (0.70 – 1.94) | 6.93% (5.02 – 8.92) | 1.85% (1.00-3.23) | 2.65% (1.65-3.86) | 2.18% (1.21-3.43) | 4.94% (3.61 – 6.52) |
| Cases reported<sup>b</sup> by date of last specimen collection<sup>c</sup> | 4,308 | 53,803 | 10,525 | 6,794 | 4,493<sup>d</sup> | 29,287 |
| Estimated cumulative infections | 48,291 (29,915 – 82,907) | 641,778 (464,896 – 826,070) | 117,389 (63,453 – 204,955) | 161,936 (100,828 – 235,877) | 47,373 (26,294 – 74,537) | 176,012 (128,624 – 232,307) |
| Estimated infections/reported cases (range)<sup>e</sup> | 11.2 (6.9 - 19.2) | 11.9 (8.6 - 15.4) | 11.2 (6 - 19.5) | 23.8 (14.8 - 34.7) | 10.5 (5.5 - 15.5) | 6.0 (4.3 - 7.8) |

<sup>a</sup> Standardized to the age and sex distribution of the counties in each region from which most specimens originated. All estimates are adjusted for test performance characteristics (specificity 99.3% (confidence interval [CI] 98.32 – 99.88%); sensitivity 96.0% (CI 89.98 – 98.89%). <sup>b</sup> COVID-19 Map by County and State. *USA Facts* https://usafacts.org/visualizations/coronavirus-covid-19-spread-map/. <sup>c</sup> Dates of last specimen collection, by site were: Washington (Puget Sound): 4/1/2020; New York Metro: 4/1/2020; South Florida: 4/10/2020; Missouri: 4/25/2020; Utah: 5/3/2020. <sup>d</sup> The number of specimens for persons aged 0-18 years was inadequate, and those <18 years were excluded from the analysis. Estimates are for adults ≥19 years. Case report data was not available by age in all cases and was estimated by publicly reported age grouping. <sup>e</sup> Estimated number of times greater for infections suggested by seroprevalence estimates compared with reported cases. Range derived from 95% confidence interval of seroprevalence estimates.

Estimates of under-ascertainment using the date 7 days prior to start of specimen collection are shown in **eTable 2**. Using these earlier dates, our point estimate for under-ascertainment was lowest in CT, where estimated infections were 8.9 times greater than reported cases as of April 19, 2020 and highest in NYC, where the estimated 641,778 infections were >1,000 times greater than the 545 cases reported by March 16, 2020. These estimates do not take into account delays in reporting results, which may have been longer earlier in the pandemic.

## Discussion

Our study estimated seroprevalence of antibodies to SARS-CoV-2 virus in six diverse geographic sites with discrete collection periods from late March through early May 2020. We found that an estimated 1.13% of persons in the Puget Sound Region of Washington State and 6.93% of those in the New York City metro region had antibodies to SARS-CoV-2 by late March. Seroprevalence varied from 1.85% in south FL in early April to 4.64% in CT by early May 2020. Our results for each site suggest that the number of infections was much greater than the number of reported cases throughout the study period; these infections likely include asymptomatic and mild infections for which health care was not sought, as well as symptomatic infections in persons who either did not seek care or on whom SARS-CoV-2 viral testing was not performed. It is possible that false positive ELISA results, especially in WA, FL, and UT, where reported cases and seroprevalence were lower, could lead to us to overestimate seroprevalence and infections. The estimates from these six regions are the first reported from a projected set of specimens from 10 U.S. states. To assess possible geographic differences and changes over time, specimens will be collected from the same areas at a variety of time points.^16^

The results of several U.S. seroprevalence studies have been released, including those conducted in Santa Clara County (CA), Idaho, Los Angeles, and New York state.^17-20^ Different assays and participant selection methods were used in each of these studies. The Santa Clara study, conducted April 3 and 4, 2020, approximately 5 weeks after the first case of community transmission of COVID-19 was detected in the San Francisco Bay area, estimated a seroprevalence rate of between 2.49% and 4.16% in a pre-print publication.^17^ The authors noted that seroprevalence estimates were largely driven by estimates of test performance characteristics, which is to be expected, particularly in a low prevalence setting. The study in New York state used sera collected approximately 3-4 weeks after our study and 8 weeks after community transmission was first identified there. Seroprevalence of 14% was estimated, with a wide variation by specific geographic location.^20^ The higher seroprevalence found in this study compared with our results may reflect later specimen collection by several weeks in that study, during a period when SARS-CoV-2 was circulating widely in New York City.

The relationship between the presence of antibodies to SARS-CoV-2 and protective immunity against future infection is currently unknown.^15^ Extrapolating these estimates to make assumptions about population immunity should be done with caution until more is known about the correlation between both the presence and duration of antibodies and protection against this novel, emerging disease.

The exact timing of the development of SARS-CoV-2-specific antibodies is variable; it is unknown when infection occurred for individuals in this study. While humoral response kinetics to SARS-CoV-2 infection are not well understood, reactive IgA, IgM, and IgG antibodies have been detected as soon as one day after symptom onset.^21^ Other studies have demonstrated that neutralizing antibodies were detected 10-15 days after onset. The median time to development of total antibody, IgM, and IgG has been estimated as 11, 12, and 14 days, respectively.^13,15^ We compared the number of estimated cases in the population based on our seroprevalence estimates with the reported cases as of the last day of specimen collection. From this comparison we estimated that for WA and NY, at least 10 times as many SARS-CoV-2 infections occurred in the most-affected counties as compared with reported cases of COVID-19 by the same time point in those geographic sites. For samples collected at later time points in other sites, we estimated that there were from 6 times as many SARS-CoV-2 infections as reported cases in CT to 23 times the number of infections as reported cases in MO. Specimen collection in CT started later than the other sites, and lower under-ascertainment estimates for CT may reflect increasing availability of testing as the pandemic progressed. Estimates of under-ascertainment are conservative; they would be many times higher if one used an earlier date to take into account infected persons who had not yet developed detectable antibodies at the time of specimen collection. Our seroprevalence estimates are more likely to reflect infections that occurred a minimum of 1 to 2 weeks prior to the specimen collection.

The study has limitations associated with the samples and with the tests used. The specimens were deidentified and originally collected for clinical purposes from persons seeking healthcare and were shared with CDC with minimal accompanying data. No data on recent symptomatic illness, underlying conditions, or possible COVID-19 exposures were available. For Lab B, in some sites it is possible that multiple specimens came from the same individual, potentially biasing results. Residual clinical specimens from screening or routine care are more likely to come from persons who require continued monitoring for chronic medical conditions despite the ongoing pandemic. These persons may not be representative of the general population, including in their healthcare seeking and social distancing behavior, immune response to infection, and disease exposure risk. Representativeness may vary as well by age group. Therefore, our seroprevalence estimates need to be confirmed by other studies, including serosurveys which use targeted sampling frames to enroll more representative populations.^22^ In addition, although overall sample size was large, there were a limited number of specimens in those <5 years of age, which limited our ability to estimate seroprevalence among young children. Furthermore, at this stage in the pandemic infections may not be evenly distributed even within these geographic sites, and seroprevalence estimates for large geographic sites (e.g., statewide for CT, MO, and UT) may not be accurate if the majority of samples come from specific areas with higher infection rates at the time of specimen collection. We also had limited geographic data on a subset of specimens from Lab B for NY and WA, which may have been drawn from a larger geographic site than those from Lab A with zip code–level data. The inclusion of some specimens from other sites of Washington and New York state, especially sites of lower seroprevalence, may lead to inaccurate seroprevalence estimates for these regions and may explain the differences in the seroprevalence estimates between Lab A and Lab B for NYC. Finally, the representation of specific geographic pockets may not be the same between the two commercial laboratories and underlying patient populations may differ between labs; therefore, combining results from Lab A and Lab B may not be valid. Follow-up serosurveys will include zip code data for all specimens.

It is possible that the ELISA may exhibit cross-reactivity with antibodies to other common human coronaviruses; therefore, some results may represent a false positive result for SARS-CoV-2, potentially leading to overestimation of the actual seroprevalence. The assay used has high specificity for SARS-CoV-2 and cross-reactivity with common coronaviruses generated results below the cut-off used for this assay.^10^ However, even with a highly specific test, the effect of false positives may be particularly marked in lower prevalence settings, including WA, UT, and FL, when specimens were collected. We did consider the performance characteristics of the ELISA when making seroprevalence estimates. Although the assay has high sensitivity (96%), it is not 100% sensitive and thus not all persons with antibodies will be detected by this assay. Finally, several early reports indicate that not all persons with SARS-CoV-2 infection (confirmed by RT-PCR testing of respiratory specimens) mount an antibody response, and antibody titers may be lower in those with milder disease.^15,23^ For these reasons, seroprevalence estimates may underestimate the proportion of persons with prior infection in any population.

Tracking population seroprevalence serially, in a variety of specific geographic sites, will inform models of transmission dynamics and policy decisions regarding the impact of social distancing and other preventive measures. We plan to conduct repeat sampling in these and other geographic sites around the United States on an ongoing basis. Seroprevalence studies such as this one will inform our understanding of the epidemiology of COVID-19. Our seroprevalence estimates suggest that at the time of specimen collection during March-early May 2020, a large majority of persons in six diverse geographic sites had not been infected with SARS-CoV-2 virus. The estimated number of infections was much greater than reported cases in all sites, potentially reflecting persons who had mild or no illness or who did not seek medical care or get tested, but who still may contribute to ongoing virus transmission in the population. Because persons do not always know if they are infected with SARS-CoV-2, the public should continue to take steps to help prevent the spread of COVID-19, such as wearing cloth face coverings when outside the home, remaining six feet apart from other people, washing hands frequently, and staying home when sick.

## Data Availability

A limited dataset will be made publicly available at a later time.

## Acknowledgements

We would like to thank LabCorp and Quest for supplying specimens. From Quest: William A. Meyer III, Larry A. Hirsch, Taylor Hwang and Janet M. Rochat. From the New York City Department of Mental Health and Hygiene: Marcelle Layton. From the Centers for Disease Control and Prevention (CDC) Molecular Pathogenesis and Immunology Research Laboratory team: Bailey Alston, Muyiwa Ategbole, Shanna Bolcen, Darbi Boulay, Peter Browning, Li Cronin, Ebenezer David, Rita Desai, Monica Epperson, Yamini Gorantla, Lily Jia, Han Li, Pete Maniatis, Jeff Martin, Kimberly Moss, Kristina Ortiz, Palak Patel, So Hee Park, Yunlong Qin, Evelene Steward-Clark, Heather Tatum, Andrew Vogan, Brianna Zellner.

Fiona Havers and Travis Lim had full access to all the data in the study and takes responsibility for the integrity of the data and the accuracy of the data analysis.

## Financial Support

This work was supported by the Centers for Disease Control and Prevention, Atlanta, GA.

## Potential conflicts of interest

The authors report no potential conflicts of interest.

## Notes

### Competing Interest Statement

The authors have declared no competing interest.

### Funding Statement

This study was funded by the Centers for Disease Control and Prevention.

### Author Declarations

This protocol underwent review by CDC human subjects research officials, who determined that the testing represented non-research activity in the setting of a public health response to the COVID-19 pandemic.

